# A Mobile Health Approach for Monitoring Hypertensive and Mental Health Conditions to Avoid Preventable Delays in Postpartum Care

**DOI:** 10.1101/2025.10.03.25337009

**Authors:** Sepideh Nikookar, Daniel Phan, Chad Robichaux, Marcia Watson, Kimberly Carroll, Banafsheh Bonnie Shoai, Marissa Platner, Natalie D. Hernandez, Sheree L. Boulet, Cheryl Franklin, Gari D. Clifford, Nasim Katebi

## Abstract

The postpartum period presents a critical window for maternal health, yet it is often characterized by limited clinical follow-up. This study evaluated the feasibility of implementing a mobile health (mHealth) system for remote monitoring of Blood Pressure (BP) and mental health among postpartum individuals. A total of 98 postpartum participants were enrolled, of whom 60 with complete and verifiable data were monitored for up to 12 weeks via the MOYO-Mom platform, with all participants self-identified as African Americans. In addition to evaluating user engagement and adherence to the study protocol, associations were examined between depressive symptoms, measured by the Edinburgh Postnatal Depression Scale (EPDS), and key variables including BP, age, and Hypertensive Disorders of Pregnancy (HDP). Of the enrolled cohort, 45 participants contributed 727 valid home BP measurements and 44 completed the EPDS at least once, providing longitudinal physiological and mental-health data for analysis.

Findings from this study indicate that lower systolic BP was significantly associated with higher EPDS scores (*p <* 0.001), suggesting a physiological link to postpartum depression. Additionally, individuals with higher depression scores and those diagnosed with HDP demonstrated lower engagement with the mHealth platform. In contrast, younger participants showed higher adherence to the study protocol. These results—grounded in participant-level data from 423 BP readings and EPDS assessments—support the feasibility of mHealth-enabled postpartum monitoring while highlighting differential engagement across risk profiles. These findings highlight the potential of mHealth tools for postpartum care and underscore the importance of designing targeted strategies to enhance engagement,

## 1. Introduction

The postpartum period is a critical window for maternal health, marked by an elevated risk of both physical and psychological complications. Alarmingly, an estimated 60% of pregnancy-related deaths occur after delivery American College of Obstetricians and Gynecologists” Presidential Task Force on Pregnancy and Heart Disease and Committee on Practice Bulletins—Obstetrics (2019); American College of Obstetricians and Gynecologists (2025). In the United States, maternal mortality remains unacceptably high, with a reported 18.6 deaths per 100,000 live births in 2023, and the highest burden observed among non-Hispanic Black women (50.3 per 100,000)Centers for Disease Control and Prevention (2024). Georgia faces an even greater challenge, with 37.9 pregnancy-related deaths per 100,000 live births between 2020 and 2022—among the highest rates in the nation Georgia Department of Public Health (2025). Global estimates similarly emphasize the ongoing burden of maternal mortality, with approximately 260,000 women dying from pregnancy- or childbirth-related causes in 2023 World Health Organization (2023). Despite these alarming statistics, postpartum care remains fragmented and limited in duration, contributing to delayed detection and management of complications.

To better understand the drivers of maternal mortality, the “Three Delays Model” provides a valuable framework for identifying where breakdowns in care occur Thaddeus and Maine (1994). The model includes:

### (i) Delay in Decision to Seek Care

One of the primary delays in addressing maternal mortality is the decision to seek care. Various sociocultural and economic factors contribute to this delay. A key challenge is women”s lack of agency, which restricts their ability to make and carry out decisions about their own health. This limited agency is compounded by barriers posed by family support and economic means, including insurance. It is also closely linked to lack of awareness, education, understanding, and health literacy, often shared by family members, caregivers, and community members, and further exacerbated by social permission and household decision-making dynamics. The low status of women in many societies further reduces their decision-making power, often leading to hesitation in seeking medical help. Negative past experiences with healthcare services can also discourage women from seeking timely help, while fatalistic attitudes or acceptance of maternal death further compound the problem.

### (ii) Delay in Reaching Healthcare Facilities

The second delay involves challenges in reaching healthcare facilities. Geographical barriers, such as the distance to health centers and hospitals, pose significant challenges, especially in rural and remote areas. Transportation issues, including limited access to affordable and reliable transportation options, further hinder timely access to care. Poor infrastructure, characterized by inadequate road networks, exacerbates these difficulties.

### (iii) Delay in Receiving Adequate Care

The third delay pertains to the provision of adequate care upon reaching a healthcare facility. Resource scarcity, including poor facilities and a lack of medical supplies, often hinders safe deliveries. Healthcare workforce challenges, such as staff shortages and inadequately trained personnel, further compromise the quality of care. Moreover, gaps in the referral system can delay the transfer of patients to higher-level facilities when needed.

Addressing these delays is essential to improving maternal health outcomes. New models of care that improve awareness, accessibility, and quality of services may help reduce preventable maternal deaths and promote more equitable health outcomes.

One of the most prevalent yet under-addressed complications in the postpartum period is postpartum depression (PPD), which affects up to 1 in 8 women in the United States Wisner et al. (2013). It is characterized by persistent feelings of sadness, anxiety, and fatigue, often accompanied by changes in sleep and appetite. While PPD has been extensively studied in psychiatric and behavioral health contexts, emerging evidence suggests relationship with physiological features, particularly blood pressure (BP). Moreover, PPD disproportionately affects low-income and minority populations, further exacerbating health disparities Stewart and Vigod (2019). The Edinburgh Postnatal Depression Scale (EPDS) is the most widely used screening tool for identifying PPD symptoms and is a central instrument in many maternal health studies Cox et al. (1987); Gibson et al. (2009).

In addition to mental health conditions like PPD, Hypertensive Disorders of Pregnancy (HDP) including gestational hypertension and preeclampsia are leading contributors to maternal morbidity and mortality. Globally, HDP affect approximately 5–10% of pregnancies and represent the second leading cause of maternal death, accounting for an estimated 14% of maternal mortality worldwide Duley (2009); Khan et al. (2006). In the United States, these conditions complicate approximately 14% of delivery hospitalizations and contribute to nearly one-quarter of severe maternal morbidities 12qaFord (2022); Centers for Disease Control and Prevention (2023); Petersen (2019); Gunderson et al. (2025). Although often considered complications of late pregnancy, the impact of HDP extends well into the postpartum period. Individuals with a history of HDP are about 50% more likely to be readmitted to the hospital or visit the emergency department than those without HDP Horwitz et al. (2021), and they face elevated long-term risks of cardiovascular disease. Moreover, HDP disproportionately affect African American women, who are three to four times more likely to die from pregnancy-related causes—including HDP—than non-Hispanic White women Tucker et al. (2007); Centers for Disease Control and Prevention (2019). Understanding how HDP intersect with mental health conditions such as PPD may offer critical insights for developing comprehensive postpartum care models.

Despite these risks, clinical follow-up during the postpartum period is often limited, with many individuals not receiving timely evaluation or support. A systematic review found that postpartum visit attendance in the U.S. is highly variable and frequently inadequate, with attendance rates as low as 50% in some populations Attanasio et al. (2022). This underscores the need for continuous monitoring strategies that integrate both physical and mental health assessments. Remote monitoring using mobile health (mHealth) monitoring systems have emerged as powerful tools to extend care beyond the clinical visits in both high- and low-resource settings. The systematic review by Knop et al. (2024) highlights the significant impact of mHealth interventions from conception to 24 months postpartum. In addition, recent research by Kumar et al. (2022) has highlighted the importance of managing postpartum hypertension effectively, emphasizing strategies such as diuretic use, remote BP monitoring through telehealth, improved transitions to primary care, and policy changes such as extended Medicaid coverage to support ongoing postpartum care. Innovations such as telemedicine models, remote assessment of ambulatory BPs, and risk prediction models using machine learning contribute to improved clinical management. Additionally, policy changes, such as postpartum Medicaid extension and emphasizing primary care transitions, may enhance long-term outcomes for women with postpartum hypertension. These interventions encompass mobile apps, SMS reminders, and telemedicine services, enhancing communication, providing timely health information, and facilitating remote monitoring of maternal health. The study by Bt Wan Mohamed Radzi et al. (2020) surveyed 819 postpartum women with obesity and depression, finding that those who used fitness apps weekly or daily experienced significant reductions in both BMI and depressive symptoms. Findings underscore the importance of personalized interventions delivered through mobile apps, including features such as symptom tracking, medication reminders, and mental health support. These tools enable women to actively manage their health during this critical postpartum period.

The study team developed the MOYO-Mom mHealth platform to investigate the relationship between postpartum depressive symptoms, BP trends, and participant engagement in a high-risk population. The platform enabled postpartum women to self-report BP readings and complete digital EPDS assessments over a 12-week period. Alongside these self-reported measures, sociodemographic and clinical data—including age, marital status, race/ethnicity, education, employment status, and HDP diagnosis—were collected. Analyses focused on understanding the interplay between psychological and physiological factors and how these associations influence adherence to the monitoring protocol. Engagement with the mHealth system was further assessed to explore how factors such as depression severity, hypertensive risk, and age affected study adherence. By combining longitudinal measurements, correlation analysis, and mixed-effects modeling, the study aims to identify barriers to sustained engagement and offer insights into maternal health patterns and the feasibility of using digital tools for continuous postpartum care.

Section 2 describes the mHealth system and study methodology; Section 3 presents the main findings; Section 4 discusses the implications of the results and limitations; and Section 5 concludes with final takeaways.

## 2. Methods

### 2.1. MOYO-Mom System Overview

This study employed a prospective observational design to monitor postpartum individuals over a 12-week period using a digital health platform. The goal was to collect longitudinal data on physiological and psychological parameters in a real-world setting.

The MOYO-Mom platform is a mobile health (mHealth) system composed of two components: a smartphone application used by participants and a secure web-based dashboard used by clinicians. It is compliant with the Health Insurance Portability and Accountability Act (HIPAA), which sets national standards for protecting sensitive patient health information. The application was installed on participants” personal smartphones and designed to facilitate structured data collection throughout the postpartum period.

Users received an OMRON 3 Series Upper Arm BP Monitor (BP7100), a device that meets international standards for automated BP monitors and has been validated for use during pregnancy in accordance with the European Society of Hypertension International Protocol (ESH-IP) STRIDE BP International Organization (2024). Participants were instructed to measure their BP at scheduled intervals, and all measurements were automatically transmitted to a secure, HIPAA-compliant cloud server. Clinical teams reviewed incoming data to ensure continuous oversight and timely follow-up.

The system also incorporated structured digital surveys that collect sociodemographic characteristics, psychosocial data, and pandemic-related stressors (e.g., impact of COVID-19), completed at predefined intervals. Detailed survey content and timelines are described in section 2.2. In addition, the app includes symptom tracking, enabling users to log clinical signs such as headaches, dizziness, or visual disturbances. When critical BP thresholds or severe symptoms were recorded, the system automatically generated alerts to both the user and the clinical team.

Communication between users and healthcare teams is facilitated through a secure interface embedded in the application, safeguarding sensitive health information while enabling responsive care. In the event of abnormal readings such as severely elevated BP or symptoms suggestive of preeclampsia the system issues urgent alerts. If the systolic BP exceeded 160 mmHg or the diastolic BP exceeded 110 mmHg, the patient was prompted to wait 15 minutes and repeat the measurement. If the BP remained elevated, an alert was triggered. These alerts were sent to clinicians via SMS and email, allowing for immediate triage and potential intervention. A view of the clinician-facing dashboard used for symptom and BP review is shown in figure 2. Users also received in-app pop-up messages with instructions, including recommendations to seek emergency care when needed. The alerting and response workflow is depicted in Figure 1.

**Figure 1.**
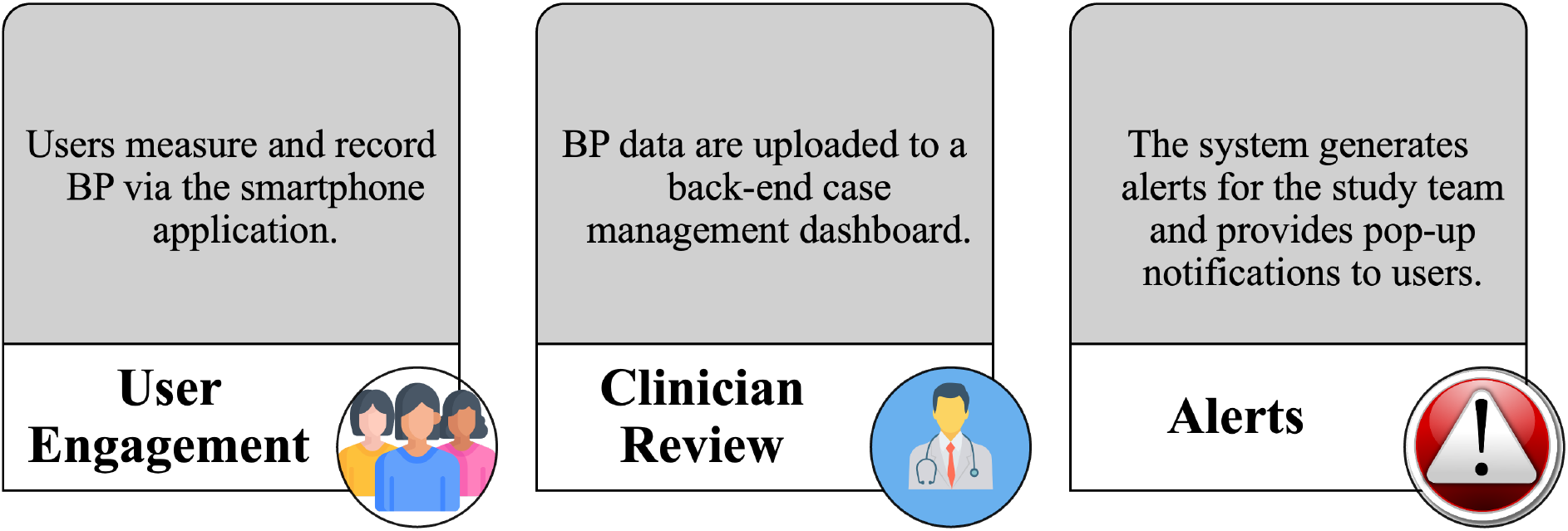
The MOYO-Mom alert system workflow.

**Figure 2.**
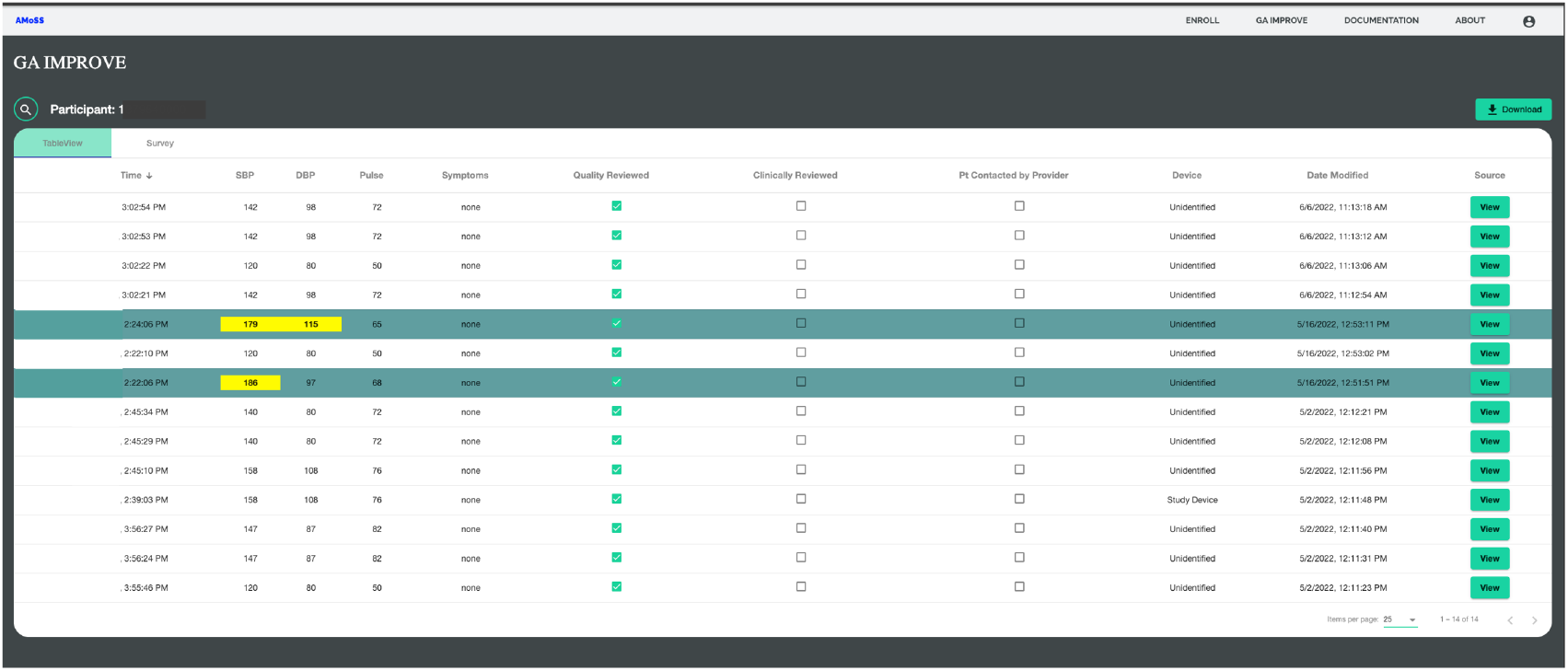
Clinician-facing dashboard within the MOYO-Mom monitoring system, built on an AWS cloud infrastructure. The dashboard displays a tabular view of participant-submitted data, including date and time of entry, SBP and DBP, pulse, and reported symptoms. Each row represents an individual reading. Columns track review and follow-up status through checkboxes for “Quality Reviewed,” “Clinically Reviewed,” and “Patient Contacted by Provider.” The interface also includes device source, modification timestamp, and a link to view full details. This tool enables clinicians to monitor BP trends in real time, identify critical values, and initiate appropriate follow-up actions.

By integrating physiological data with real-time self-reporting and automated alerts, MOYO-Mom offers a proactive and personalized model for postpartum care. This system enhances early detection of complications, supports timely clinical response, and facilitates patient education and empowerment in the recovery period.

Figure 3 shows a screenshot of the MOYO-Mom mobile interface. The app features a user-friendly design where participants can log symptoms, view recent BP entries, and complete digital surveys. Reminders were embedded in the system to help users stay engaged with the monitoring protocol, contributing to consistent data collection and timely alerts.

**Figure 3.**
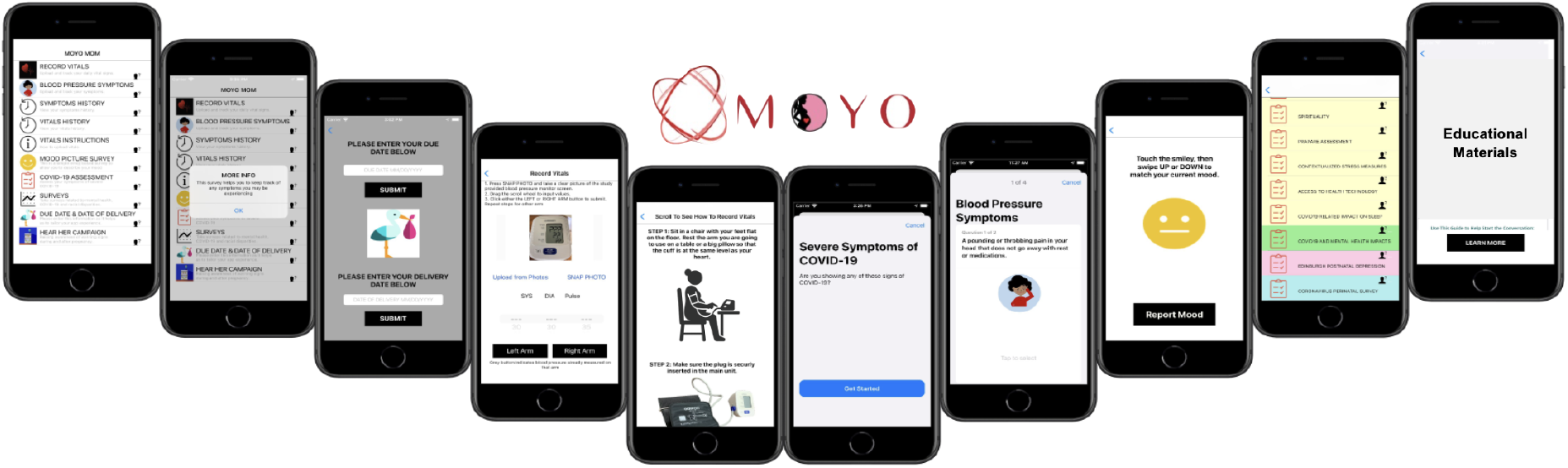
Participant-facing interface of the MOYO-Mom mobile application. The app supports structured postpartum monitoring and includes features for recording BP, reporting symptoms, completing digital surveys (e.g., EPDS, COVID-19 impact, mood assessments), and accessing educational materials. The screenshots illustrate key functionalities such as vitals entry, symptom tracking, survey selection, and health guidance prompts, designed to support user engagement and self-monitoring during the postpartum period.

### 2.2. Study Protocol

The study protocol was designed to support continuous monitoring, participant engagement and holistic assessment of physical and mental health over a 12-week postpartum period. This study was approved by the Morehouse School of Medicine Institutional Review Board (MSM IRB 1800529-32). Participants were enrolled at clinical sites prior to delivery, where they received training and study materials. Participants progress through sequential stages, each designed to support data collection, symptom surveillance, and timely intervention.

Upon enrollment, participants received a study kit and underwent structured training on using the MOYO-Mom application. Surveys administered at enrollment encompass a broad range of psychosocial and behavioral domains, including coping strategies (COPE), spirituality, social determinants of health (PRAPARE), contextualized stress, access to health technology and the impact of COVID-19 on sleep and mental health. This phase includes a total of 164 questions aimed at capturing baseline psychosocial and contextual factors.

Participants were instructed to document delivery-related information within 48 hours postpartum. During the first postpartum week, BP is monitored twice daily, with a minimum of 7 readings required. No surveys were administered during this phase.

From week 1 to 3 postpartum, BP monitoring continued twice daily, with a minimum of 14 readings. Participants completed the 10-question EPDS during this stage to assess depressive symptoms.

Between weeks 3 and 6 postpartum, daily BP monitoring was required, with a minimum of 6 readings. Participants completed three surveys: a 10-question depression scale, a 56-question COPE inventory, and a 13-question COVID mental health impact assessment, totaling 92 questions.

From week 6 to 12 postpartum, participants transitioned to weekly BP monitoring with at least 6 readings. The COPE survey was repeated during this phase, comprising 56 questions.

The protocol design aimed to maintain participant compliance while enabling high-resolution data capture across physiological and psychological domains. A detailed overview of study components, including monitoring frequency, and surveys, is provided in Table 1. Participants were compensated for completing surveys and monitoring tasks.

**Table 1.** Timeline of postpartum study activities, including BP monitoring and survey administration from enrollment to 12 weeks postpartum.

| Timing | Action | Surveys |
| --- | --- | --- |
| Enrollment | App training | 1. COPE (14)<br>2. Spirituality (32)<br>3. PRAPARE (21)<br>4. Contextualized Stress (60)<br>5. Tech Access (16)<br>6. COVID Sleep (8)<br>7. COVID Mental Health (13) |
| Delivery | Delivery documentation<br>(within 48 hrs) | None |
| 1 Week<br>Postpartum | BP monitoring 2x/day<br>($\geq 7$ readings) | None |
| 3 Week<br>Postpartum | BP monitoring 2x/day<br>($\geq 14$ readings) | 1. Depression (10) |
| 6 Week<br>Postpartum | BP monitoring 1x/day<br>($\geq 6$ readings) | 1. Depression (10)<br>2. COPE (56)<br>3. COVID Mental Health (13) |
| 12 Week<br>Postpartum | BP monitoring 1x/week<br>($\geq 6$ readings) | 1. COPE (56) |

### 2.3. Data Collection

Participants 18 years or older were recruited during prenatal care at Grady Memorial Hospital and community clinics in Atlanta, GA, USA. Figure 4 presents a summary of participant distribution across these locations, providing insight into the geographic diversity of the cohort and site-level engagement patterns.

**Figure 4.**
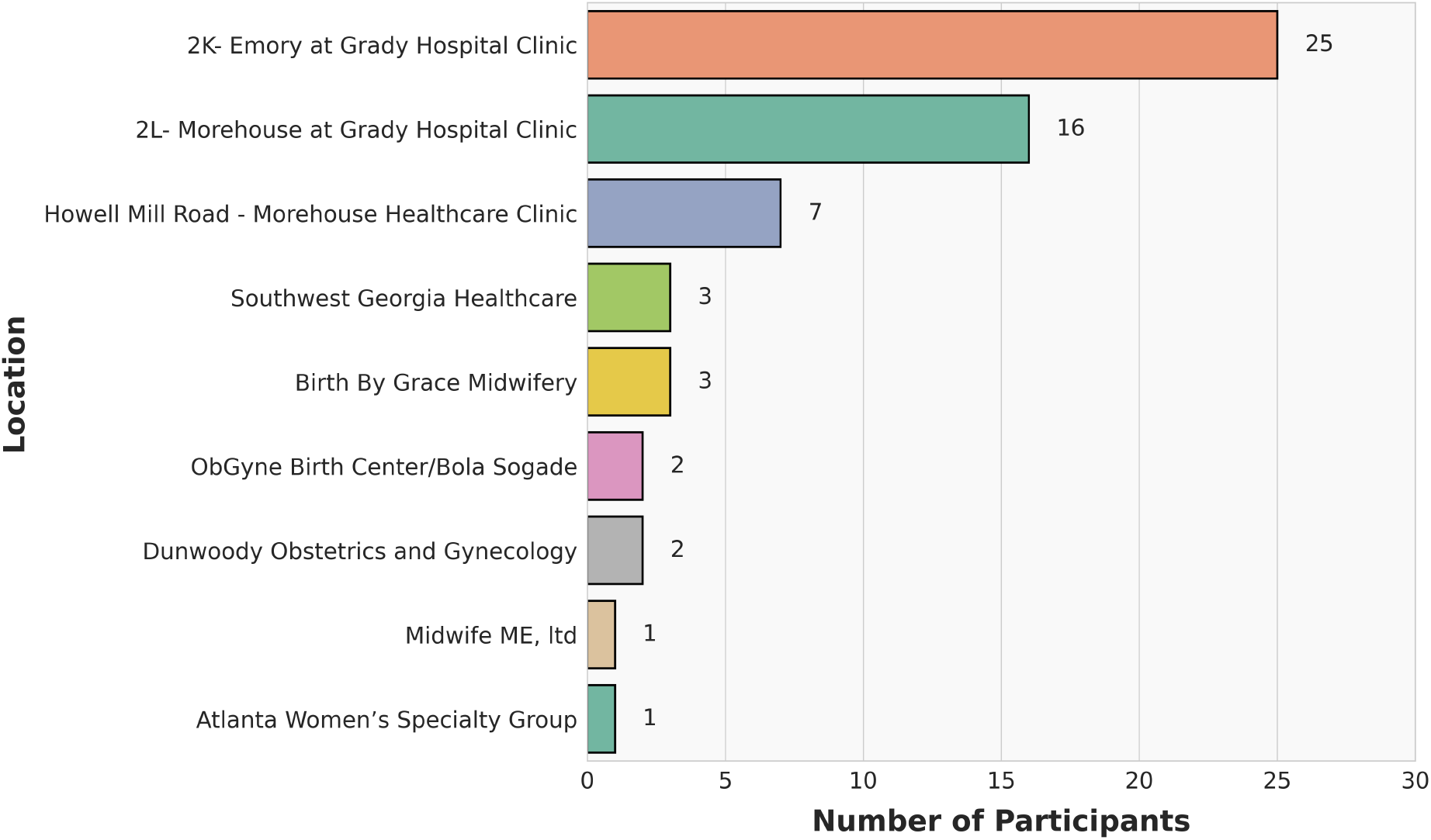
Distribution of participants across clinical recruitment sites.

The availability and completeness of data collected across key domains—including sociodemographic characteristics, BP monitoring, and clinical outcomes—are illustrated in Figure 5, which outlines the flow of participant inclusion based on data availability.

**Figure 5.**
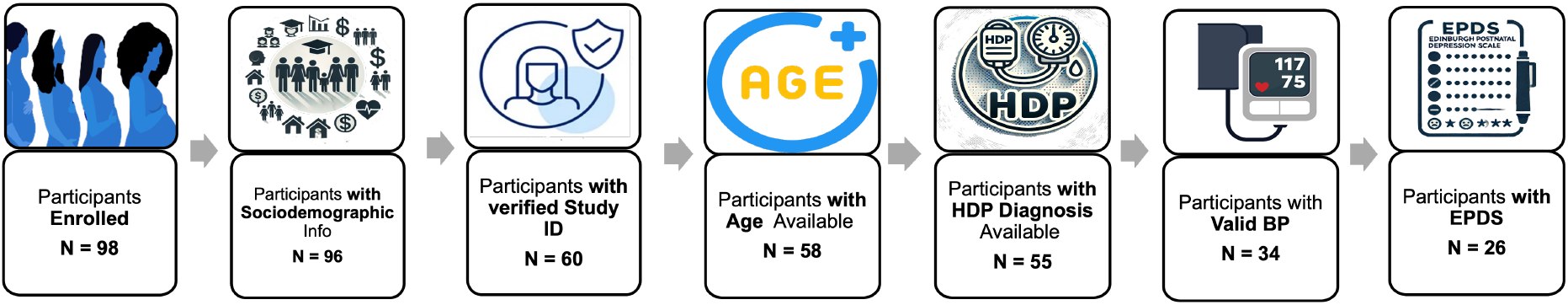
Flow diagram of participant inclusion based on data availability across key variables.

Around 98 participants were initially enrolled in the study. Sociodemographic data were available for 96 individuals; however, due to challenges in mapping these records to verified study IDs, only 60 participants could be reliably included in the final analytic dataset. Among these 60 participants, age information was available for 58, and HDP diagnoses were available for 55. Sociodemographic characteristics data were self-reported, while Age and HDP diagnoses were ascertained from medical records. These clinical and demographic variables were incorporated into subsequent analyses.

The next sections summarize the availability of data related to sociodemographic characteristics, BP monitoring, and depressive symptom screening using the EPDS.

#### 2.3.1. Sociodemographic

Sociodemographic data were available for all 60 participants and are summarized in Table 2. The cohort primarily comprised African American/Black participants (100%), with 8.3% identifying as Hispanic. Most participants (90%) were under the age of 35, and half had completed a high school diploma or GED. Approximately 70% reported having employment in 2020–2021. Regarding unmet social needs, food, utilities, and healthcare were the most frequently reported. Approximately 18.33% of participants reported food insecurity, 8.33% did not have access to a phone, 21.67% lacked reliable utilities, and 18.33% reported limited access to healthcare. Relationship status, annual household income, insurance coverage, and household size also varied, highlighting a diverse population with varying social and economic backgrounds.

**Table 2.** Sociodemographic Characteristics of Participants (N = 60)

| Variable | N (%) |  |  |
| --- | --- | --- | --- |
| **Race/Ethnicity** |  |  |  |
| African American/Black | 60 (100) |  |  |
| **Hispanic Ethnicity** |  |  |  |
| Yes | 5 (8.33) |  |  |
| No | 52 (86.67) |  |  |
| No Answer | 3 (5.0) |  |  |
| **Mother's Age** |  |  |  |
| ≥ 35 years | 6 (10.0) |  |  |
| < 35 years | 54 (90.0) |  |  |
| **Level of Education** |  |  |  |
| Did not finish high school | 8 (13.33) |  |  |
| High school diploma/GED | 30 (50.0) |  |  |
| Associate's degree | 5 (8.33) |  |  |
| Bachelor's degree | 10 (16.67) |  |  |
| Master's degree | 4 (6.67) |  |  |
| No Answer | 3 (5.0) |  |  |
| **Employment in 2020–2021** |  |  |  |
| Yes | 42 (70.0) |  |  |
| No | 15 (25.0) |  |  |
| No Answer | 3 (5.0) |  |  |
| **Reported Social Needs** |  |  |  |
| Food | 11 (18.33) |  |  |
| Phone | 5 (8.33) |  |  |
| Utilities | 13 (21.67) |  |  |
| Healthcare | 11 (18.33) |  |  |
|  |  | **Variable** | **N (%)** |
|  |  | **Relationship Status** |  |
|  |  | Single | 27 (45.0) |
|  |  | Living with Partner | 15 (25.0) |
|  |  | Married | 11 (18.33) |
|  |  | Not Living Together | 4 (6.67) |
|  |  | No Answer | 3 (5.0) |
|  |  | **Annual Household Income** |  |
| | | < \$9,000 | 19 (31.67) |
| | | \$9k–\$19,999 | 7 (11.67) |
| | | \$20k–\$29,999 | 8 (13.33) |
| | | ≥ \$30,000 | 22 (36.67) |
|  |  | No Answer | 4 (6.67) |
|  |  | **Insurance Type** |  |
|  |  | Medicaid | 45 (75.0) |
|  |  | Private | 9 (15.0) |
|  |  | No Insurance | 3 (5.0) |
|  |  | No Answer | 3 (5.0) |
|  |  | **Household Size** |  |
|  |  | 2 or fewer | 20 (33.33) |
|  |  | More than 2 | 40 (66.67) |

#### 2.3.2. Blood Pressure

Out of the 60 participants who provided sociodemographic information, 40 individuals submitted a total of 716 self-measured BP readings with delivery dates available. One participant was excluded from the analysis due to providing an unusually high number of late BP recordings (n = 238), which were submitted between two and four months postpartum—beyond the study”s monitoring window. In addition, participants who only submitted BP readings before delivery, without any postpartum values, were excluded to maintain consistency with the postpartum focus of the study.

To ensure analytic consistency, only BP data recorded from one month prior to delivery through three months postpartum were retained. Following these exclusions, the final analytic sample included 34 participants and a total of 423 valid BP measurements.

To characterize overall BP levels in the cohort, Figure 6 displays the distribution of SBP and DBP measurements categorized into four clinical ranges including Normotensive, Elevated BP, Hypertension Stage 2 and Severe Hypertension.

**Figure 6.**
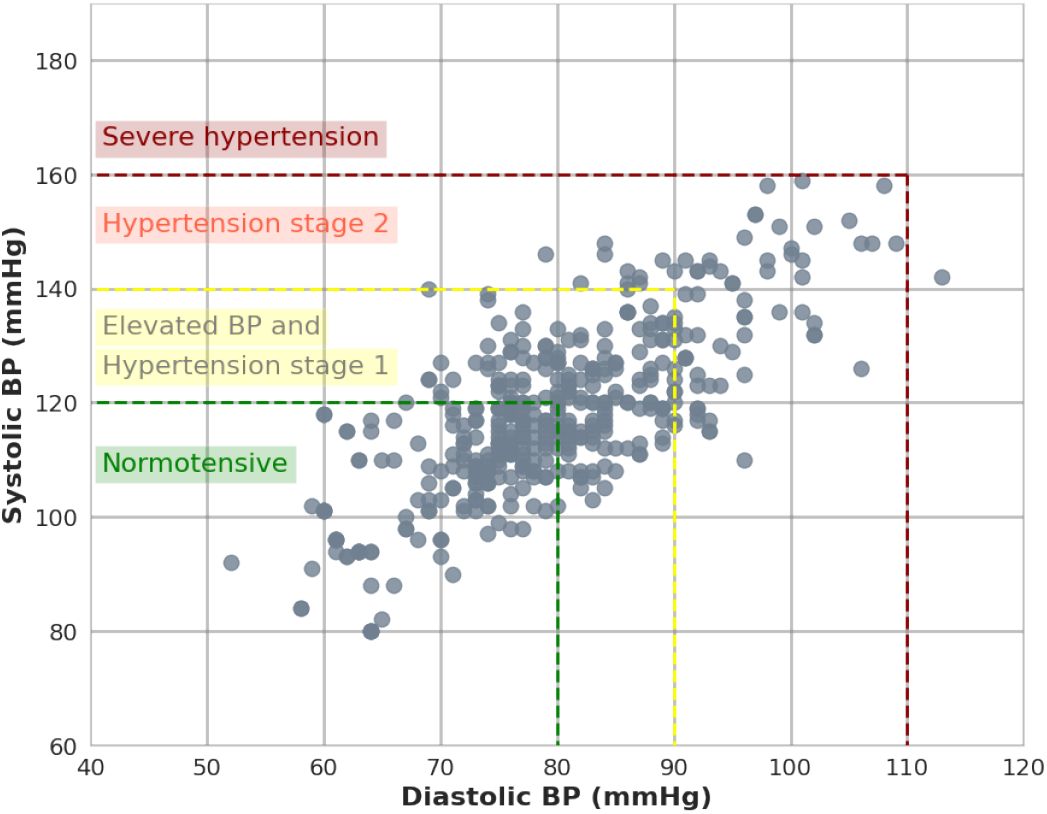
Distribution of SBP and DBP.

To explore the timing of BP measurements within the 24-hour day, we analyzed the distribution of recording times. As shown in Figure 7, the majority of BP readings were recorded during the evening, particularly after 9 PM. This trend may be attributed to the postpartum context, as this time period may offer fewer caregiving interruptions.

**Figure 7.**
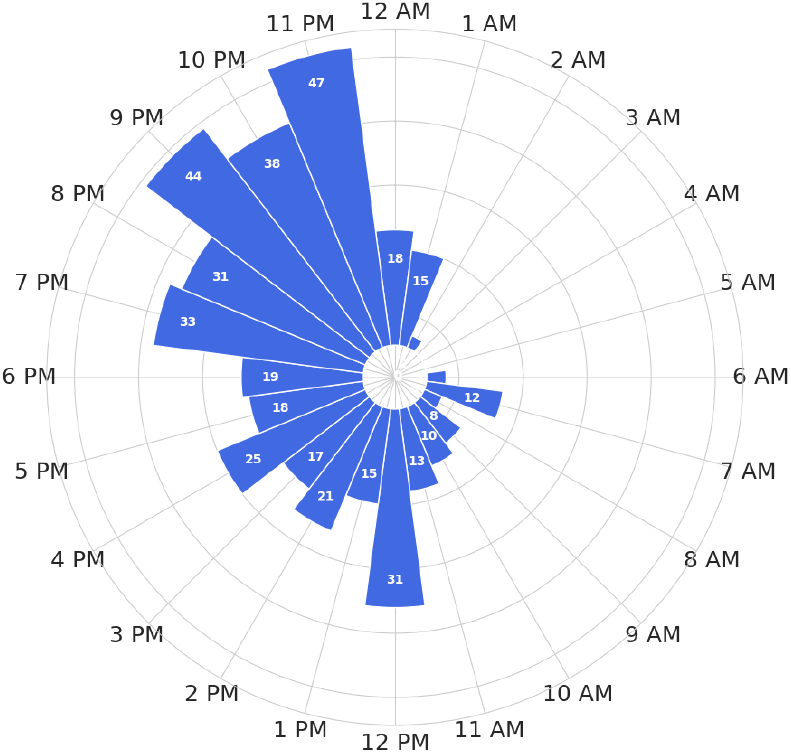
Time-of-day distribution of recorded BP readings.

To assess whether time of day influenced BP values, average SBP and DBP readings were compared between morning (AM) and evening (PM) time periods. Most participants submitted BP measurements exclusively during either the AM or PM period, rather than across both time windows. Therefore, a paired analysis was conducted only among the subset of participants who recorded readings in both the AM and PM. As shown in Figure 8, no statistically significant differences were observed in mean SBP (p = 0.11) or DBP (p = 0.38) BP values between AM and PM recordings. Given the lack of significant variation by time of day, all BP measurements were treated equivalently in subsequent analyses, regardless of the time they were recorded.

**Figure 8.**
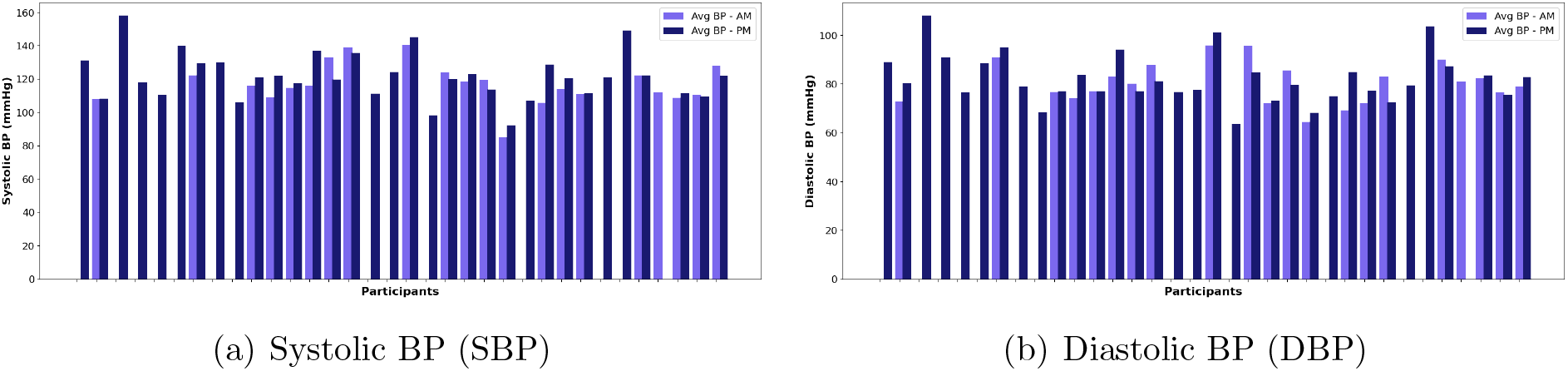
Comparison of daytime and nighttime BP values.

#### 2.3.3. Edinburgh Postnatal Depression Scale (EPDS)

The EPDS was a key component of the digital survey set administered to participants throughout the study period. Among the 34 participants who met the inclusion criteria for BP analysis, 26 completed the EPDS survey at least once. As a validated 10-item instrument, the EPDS is widely used to screen for symptoms of postpartum depression.

Figure 9 displays the distribution of EPDS scores across participants. Scores were categorized into four levels of depression severity: scores below 6 indicated minimal or no depression, scores from 7 to 13 reflected mild depression, scores between 14 and 19 suggested moderate depression, and scores above 19 indicated severe depressive symptoms McCabe-Beane et al. (2016). This stratification provided insight into the varying degrees of psychological distress experienced during the postpartum period.

**Figure 9.**
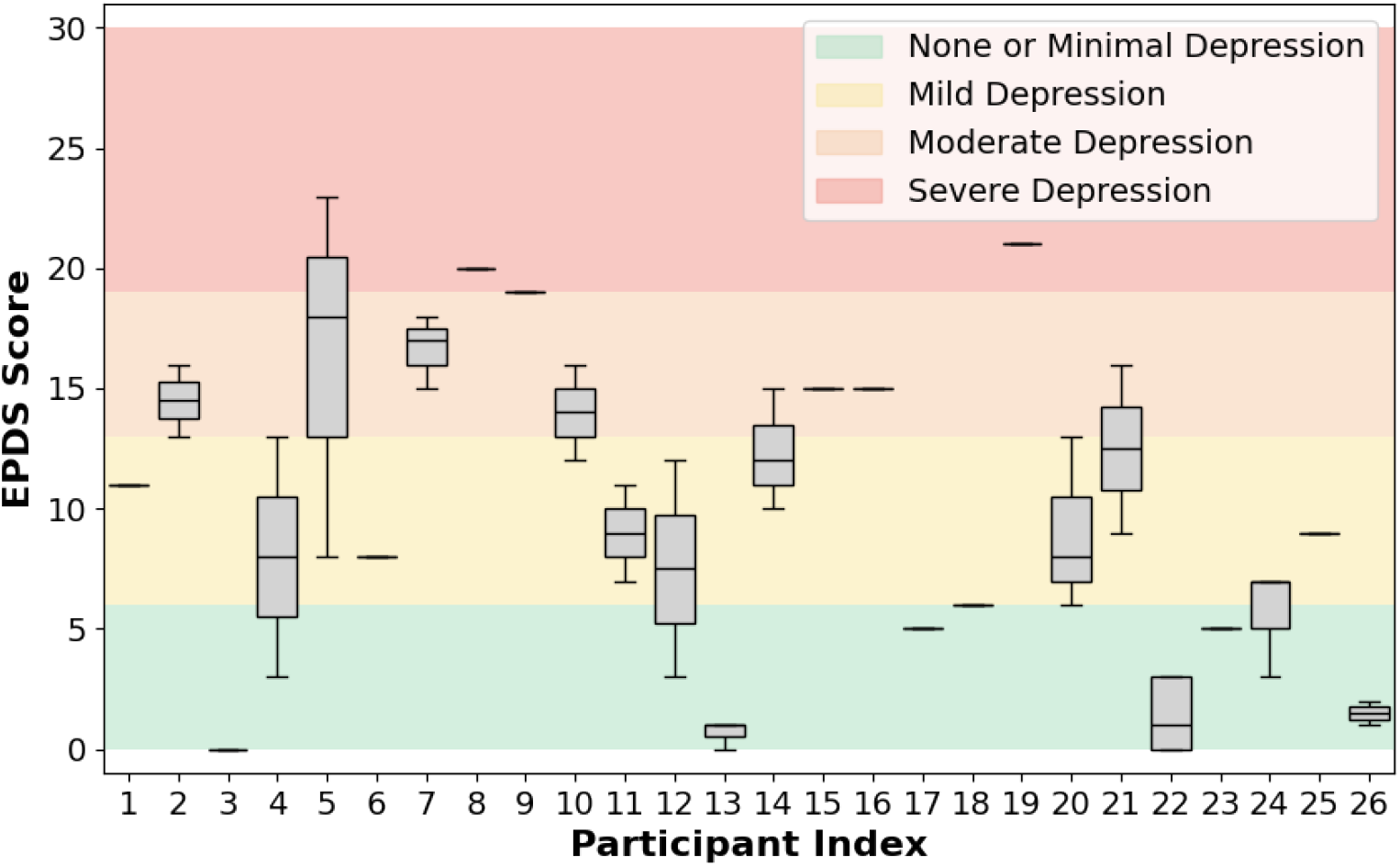
Distribution of EPDS scores among participants.

EPDS scores were analyzed in conjunction with other key variables, including BP trends, age, and HDP diagnosis. Participants with a median EPDS score of 13 or higher were classified as being at high risk for postpartum depression and were examined as a subgroup in subsequent analyses.

#### 2.3.4. Involvement Score

The involvement score is a composite measure developed to quantify participant engagement throughout the 12-week postpartum monitoring period. It reflects adherence of participation in two critical study activities: BP monitoring and completion of the EPDS survey.

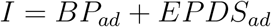

This score focuses solely on adherence to the study protocol, defined as follows:

- **Adherence components (**(.)_*ad*_): Calculated by evaluating whether the participant met minimum submission requirements during each predefined time window. Readings or surveys submitted outside of these windows were not credited in this component, even if they exceeded the required number.

The involvement score serves as an important indicator of both the completeness and quality of participant engagement. It supports unbiased analysis by accounting for variability in participation and enables assessment of the feasibility and acceptability of digital health interventions in the postpartum setting. By quantifying how consistently participants followed study protocols, this score ensures that the subsequent analyses of clinical and behavioral outcomes are grounded in reliable, representative data.

To illustrate engagement patterns across the cohort, Figure 10 presents a histogram of involvement scores for all participants. The score is normalized such that a value of 1 represents the highest possible adherence—indicating that the participant followed the study protocol exactly across all required time windows.

**Figure 10.**
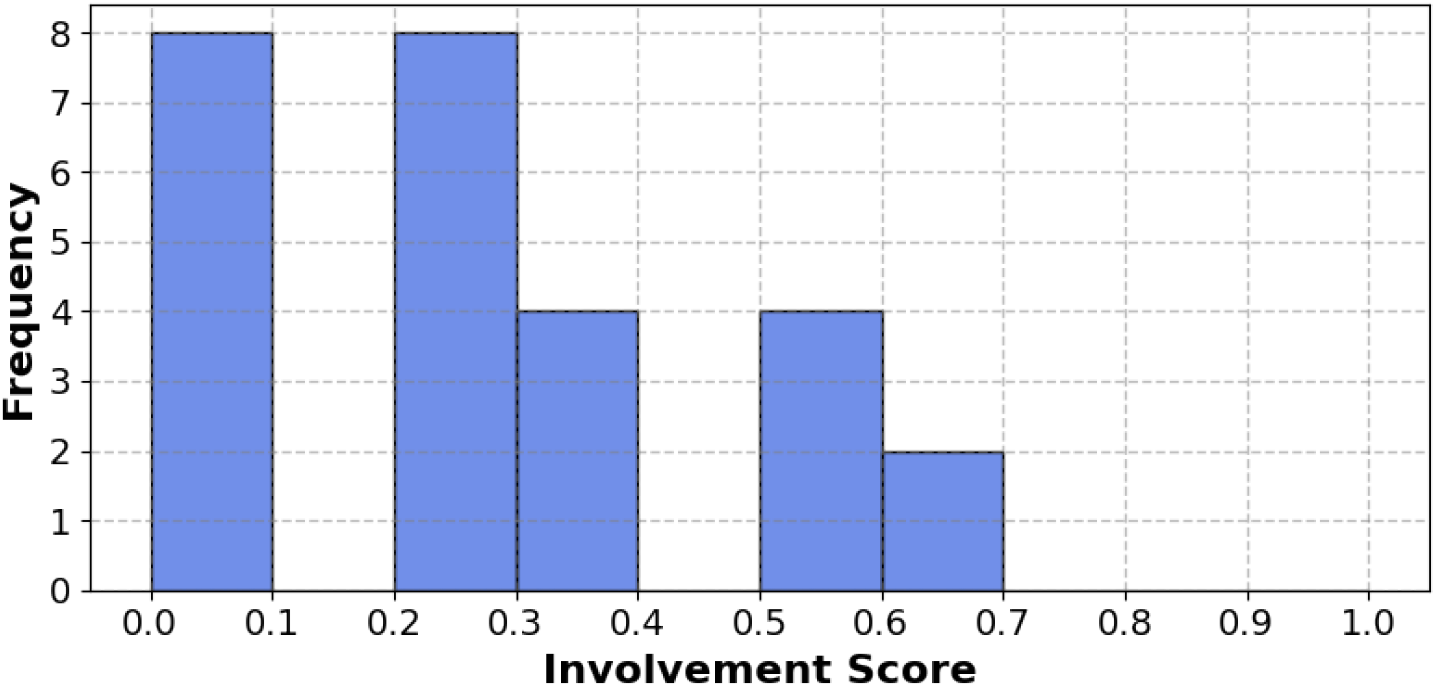
Distribution of involvement scores across study participants.

## 3. Results

### 3.1. Correlation Analysis

To examine the relationships among physiological, psychological, and clinical variables, a Pearson correlation analysis was conducted across five key features: SBP, DBP, age, HDP diagnosis, and depressive symptoms measured by EPDS. These variables were selected to capture both primary study outcomes (BP and depressive symptoms) and relevant demographic or clinical covariates (age, HDP).

For each participant, SBP and DBP values were averaged over the monitoring period, while the EPDS score was summarized using the median of all completed surveys. HDP status was coded as a binary variable. This aggregation ensured a consistent, participant-level summary for correlation analysis. The correlation results are presented in Table 3.

**Table 3.** Correlation coefficients for variables in the postpartum cohort. Pearson correlations are shown for continuous–continuous pairs. For HDP point-biserial correlations were computed.

|  | SBP | DBP | EPDS | Age | HDP |
| --- | --- | --- | --- | --- | --- |
| SBP | 1.00 | 0.89 | -0.38 | 0.05 | 0.38 |
| DBP | 0.89 | 1.00 | -0.35 | 0.03 | 0.35 |
| EPDS | -0.38 | -0.35 | 1.00 | -0.29 | 0.19 |
| Age | 0.05 | 0.03 | -0.29 | 1.00 | -0.23 |
| HDP | 0.38 | 0.35 | 0.19 | -0.23 | 1.00 |

The correlation matrix shows a strong positive correlation between SBP and DBP (*r* = 0.89) as expected. Moderate positive correlations were also observed between HDP and both SBP (*r* = 0.38) and DBP (*r* = 0.35). EPDS was moderately negatively correlated with SBP (*r* = −0.38), DBP (*r* = −0.35), and age (*r* = −0.29) and a weak positive correlation with HDP (*r* = 0.19). However, these correlations did not reach conventional statistical significance (*p >* 0.05), indicating only suggestive trends that warrant further investigation.

Building on these findings, additional analyses were conducted to more closely examine the specific association between depressive symptoms and BP levels.

Previous research has shown that individuals with clinical depression often exhibit lower systolic BP Licht et al. (2009), although this pattern has not been widely studied in the postpartum population.

### 3.2. Mixed-Effects Modeling

Mixed-effects models are particularly well-suited for longitudinal and hierarchical data where repeated measurements are nested within individual subjects. These models allow for both fixed effects—capturing population-level influences—and random effects—accounting for individual-level variability Pinheiro and Bates (2000). In this study, mixed-effects models were applied to examine how key clinical and demographic variables predict two outcomes: participant involvement and BP levels, while accounting for participant-specific variation.

#### 3.2.1. Predictors of Participant Involvement

To assess the relationship between participant characteristics and engagement, a mixed-effects linear model was fitted with the Involvement Score as the outcome variable. Because SBP and DBP were longitudinally collected predictor in the dataset, each measure was decomposed into two components: a within-person component (deviation from the participant”s own average across the study) and a between-person component (the participant”s mean level across the study). This separation avoids conflating short-term within-person fluctuations with stable between-person differences. When SBP and DBP are modeled only as raw longitudinal values, the relatively small within-person variation dominates and obscures the contribution of between-person differences. By explicitly modeling both components, it becomes possible to test whether day-to-day BP deviations or overall BP levels are associated with engagement. Fixed effects therefore included SBP-within, DBP-within, SBP-between, DBP-between, EPDS score, age, and HDP, while a random intercept was included for each participant to account for repeated measures and intra-individual variability.

Key findings are summarized in Table 4. The model identified significant negative associations between involvement score and EPDS (*p <* 0.001), HDP (*p* = 0.011), and age (*p* = 0.015), indicating that participants with greater depressive symptoms, hypertensive complications during pregnancy, or older age were less engaged with the study. Short-term within-person fluctuations in SBP or DBP were not predictive of engagement (both *p* = 1.00). However, higher between-person DBP (participants” average DBP across the study) was positively associated with involvement (*p* = 0.024), whereas between-person SBP was not significantly associated (*p* = 0.152).

**Table 4.** Mixed-effects model results for Involvement Score, with SBP and DBP decomposed into within- and between-person effects.

| Predictor | Regression Coefficient | Std. Error | 95% CI |
| --- | --- | --- | --- |
| Intercept | 0.405 | 0.134 | [0.143, 0.667] |
| SBP-within | 0 | 0 | [0, 0] |
| DBP-within | 0 | 0 | [0, 0] |
| SBP_between | −0.002 | 0.002 | [−0.006, 0.001] |
| DBP_between | 0.005 | 0.002 | [0.001, 0.010] |
| EPDS | −0.011 | 0.002 | [−0.015, −0.006] |
| Age | −0.004 | 0.002 | [−0.007, −0.001] |
| HDP | −0.066 | 0.026 | [−0.117, −0.015] |

These findings highlight the importance of psychosocial and clinical fac-tors—particularly depressive symptoms, HDP, and age—as well as stable between-person differences in DBP—over short-term physiological fluctuations in shaping participant engagement in postpartum remote monitoring.

#### 3.2.2. Predictors of Participant Depression

To investigate factors associated with postpartum depressive symptoms, a mixed-effects linear model was fitted using EPDS as the dependent variable. Because the focus was on between-person differences, participant-level averages of SBP and DBP (SBP-between and DBP-between) were used instead of raw longitudinal values. EPDS was also modeled at the between-person level by including each participant”s median EPDS across visits. Fixed effects therefore included SBP-between, DBP-between, involvement score, age, and HDP diagnosis. A random intercept for participant ID was included to account for repeated measurements within individuals.

As shown in Table 5, the model revealed several significant predictors of depressive symptom severity. Higher involvement scores were strongly associated with lower EPDS values (*p <* 0.001), indicating that participants with greater engagement in self-monitoring and survey completion reported fewer depressive symptoms. Higher between-person SBP was also associated with lower EPDS (*p <* 0.001), suggesting that participants with higher average systolic blood pressure across the study period tended to report fewer depressive symptoms. Age showed a similar negative association (*p <* 0.001), consistent with younger participants reporting more severe depressive symptoms. In contrast, between-person DBP was not significantly related to EPDS (*p* = 0.862). HDP diagnosis was positively associated with depressive symptoms (*p <* 0.001), indicating that participants with a history of hypertensive disorders in pregnancy experienced higher EPDS scores.

**Table 5.** Mixed-effects model results for EPDS Score.

| Predictor | Regression Coefficient | Std. Error | 95% CI |
| --- | --- | --- | --- |
| Intercept | 39.04 | 2.65 | [33.85, 44.23] |
| SBP_between | −0.18 | 0.04 | [−0.27, −0.10] |
| DBP_between | −0.01 | 0.06 | [−0.13, 0.11] |
| Involvement | −6.92 | 1.41 | [−9.67, −4.16] |
| Age | −0.18 | 0.04 | [−0.26, −0.10] |
| HDP | 3.29 | 0.64 | [2.03, 4.55] |

These findings highlight the protective role of participant engagement, older age, and higher between-person SBP in relation to postpartum mental health, while also pointing to hypertensive disorders in pregnancy as an important risk factor. Together, these results underscore the need for targeted support for younger individuals and those with a history of HDP, who may be at heightened risk for postpartum depression.

## 4. Discussion

This study examined the interplay between postpartum depressive symptoms, BP patterns, HDP, and participant engagement using a mHealth monitoring system. Leveraging longitudinal data and mixed-effects modeling, the analysis revealed important insights into the psychological, physiological, and behavioral dimensions of postpartum health.

A key finding was the moderate negative correlation between depressive symptoms and both SBP and DBP. Higher EPDS scores were associated with lower BP levels, a relationship consistent with prior literature Licht et al. (2009); Hildrum et al. (2008) demonstrating that depression can be linked to reduced blood pressure. While this phenomenon has been documented in non-postpartum populations—such as Licht et al. (2009)—this study provides evidence that the same pattern may persist during the postpartum period, when both psychological and cardiovascular systems are undergoing significant transitions.

In line with established literature August et al. (2017); Brown et al. (2013), HDP were significantly associated with elevated postpartum BP levels. This finding reinforces the long-term cardiovascular burden posed by HDP Timpka et al. (2016), and highlights the importance of continued BP monitoring even after delivery. Notably, participants with HDP not only exhibited higher BP but also demonstrated significantly lower levels of study engagement, indicating potential barriers to adherence among this high-risk group.

To quantify engagement, an involvement score was introduced that captures both the frequency and protocol adherence of BP and EPDS reporting. Mixed-effects modeling revealed that greater depressive symptom severity and a history of HDP were both significantly associated with lower involvement scores. These results suggest that psychosocial and clinical vulnerabilities may reduce the likelihood of consistent participation in postpartum digital health monitoring. While younger participants also tended to have lower involvement, this association did not reach statistical significance. Interestingly, BP levels themselves were not predictive of engagement, emphasizing the importance of considering mental and obstetric history over physiological indicators alone when designing remote monitoring systems.

The findings from this study have several clinical implications. First, they underscore the value of integrating mental health screening into postpartum monitoring protocols. Second, they highlight the need for personalized engagement strategies, particularly for individuals with HDP or elevated EPDS scores. These could include adaptive survey schedules, additional outreach, and tailored approaches such as culturally sensitive messaging or the use of patient navigators to support engagement and adherence. Finally, this work demonstrates the feasibility and utility of using mHealth systems to collect multidimensional postpartum data in real time, which could enhance early detection of complications and promote more proactive care models.

This study is not without limitations. The relatively small sample size and single-system context may limit generalizability, and selection bias is possible given that participation required technological access and literacy. While the involvement score offers a useful quantitative framework, it does not capture qualitative aspects of user experience or burden. Additionally, as an observational study, causality cannot be inferred from the associations observed, and unmeasured confounding factors such as childcare support, comorbidities, or socioeconomic stressors may influence both depressive symptoms and engagement.

Future research should build on these findings by validating the involvement score in larger and more diverse cohorts, incorporating qualitative data to capture participant perspectives, and testing targeted interventions to improve engagement in high-risk subgroups. Longitudinal analyses beyond the 12-week postpartum period may also help to clarify the evolving relationship between depression, hypertension, and digital health engagement in the months following childbirth.

## 5. Conclusion

This study demonstrates the utility of mHealth platforms for integrated postpartum monitoring, highlighting important associations between depressive symptoms, HDP, BP trends, and participant engagement. Data collected through a HIPAA-compliant remote monitoring system revealed that higher EPDS scores and a history of HDP were significantly associated with lower involvement in postpartum care, while BP values alone were not predictive of engagement. Additionally, depressive symptoms showed moderate inverse correlations with both SBP and DBP, consistent with findings from prior research in non-postpartum populations.

The use of real-time data capture, automated alerts, and mixed-effects modeling provided valuable insights into the complex interplay between mental and physiological health during the postpartum period. These findings support the potential of mHealth technologies to facilitate continuous monitoring, identify barriers to engagement, and inform the development of targeted strategies to improve maternal health outcomes and promote health equity.

## Data Availability

Data access for research will require IRB approval, HIPAA-compliant security documentation, and the signing of an institutional data use agreement.

## Acknowledgments

Research reported in this publication was supported by the Imagine, Innovate and Impact (I3) Funds from the Emory School of Medicine to GC and SB and through the Georgia IMPROVE on Maternal Health, funded by NIH National Center for Advancing Translational Sciences (NCATS) as an Administrative Supplement to the Georgia Clinical and Translational Alliance, grant number UL1-TR002378 to GC, SB and CF. The content is solely the responsibility of the authors and does not necessarily represent the official views of the National Institutes of Health. NK is partially funded by a PREHS-SEED award grant K12ESO33593. We would also like to acknowledge the support of the Grady Health System, Atlanta, Georgia in conducting this research. The funders had no role in study design, data collection and analysis, decision to publish, or preparation of the manuscript.

